# Cumulative cefepime exposure in cancer patients is associated with an increased risk of ertapenem non-susceptible, meropenem susceptible Enterobacterales bacteremia

**DOI:** 10.1101/2025.09.05.25335002

**Authors:** Alex V. Stabler, William C. Shropshire, Allyson Young, Chun Feng, Hyunsoo Hwang, Samuel A. Shelburne, Nancy N. Vuong, Jovan Borjan

## Abstract

Carbapenem resistance in non-carbapenemase producing Enterobacterales can display susceptibility discordance (e.g., ertapenem-non-susceptible, meropenem-susceptible), often in patients with prior antimicrobial use. We aim to characterize risk factors for carbapenem susceptibility discordant Enterobacterales (CSD-E) bloodstream infections in immunocompromised patients. We performed a retrospective, single-center study comparing adults with Enterobacterales BSI who developed subsequent carbapenem susceptibility concordant (CSC;meropenem and ertapenem susceptible) or CSD-E BSI with the same organism within one year. Patients who developed subsequent CSD-E BSI were matched to those with repeat CSC-E BSI. Logistic regression models evaluated CSD-E risk factors. Time-varying covariate Cox proportional hazards models evaluated time-dependent changes in antibiotic exposure when estimating the association between antibiotic use and CSD-E. Comparative genomics of available paired whole-genome sequencing (WGS) data was performed to assess genetic relatedness and antimicrobial resistance (AMR) gene content. Beta-lactam survival mechanisms (BLSM) were assessed via Tolerance Disk Test (TDTest) and Population Analysis Profiling (PAP). We evaluated 829 patients with Enterobacterales BSI. Repeat BSI was CSC-E in 81 patients (9.8%) and CSD-E in 14 patients (1.7%). Matching provided 43 CSC-E controls and 14 CSD-E cases. Univariate analysis revealed any ceftazidime-avibactam (CZA) exposure was associated with developing CSD-E BSI (odds ratio[OR],7.41;p=0.01). Time-varying covariate Cox regression identified each additional day of cefepime is associated with CSD-E BSI development (hazard ratio[HR],1.055;p=0.016). Genomic data revealed beta-lactamase amplification and outer membrane porin mutations as potential CSD-E development mechanisms. CSD-E BSI risk factors in immunocompromised patients may include any CZA exposure and cumulative cefepime exposure. Further studies are warranted to validate CSD-E risk factors.

## Introduction

Gram-negative bacteria are pathogens that contribute to increasing morbidity, mortality, and antimicrobial resistance [1]. Carbapenems are broad-spectrum antibiotics that can treat extended-spectrum beta-lactamase (ESBL) producing bacteria, but resistance mechanisms are increasing, rendering carbapenems less effective [2]. Carbapenem-resistant Enterobacterales (CRE) are defined as Enterobacterales (e.g., *Escherichia coli*) that test resistant to at least one carbapenem *or* produce a carbapenemase (CP CRE), such as *Klebsiella pneumoniae* carbapenemase (KPC) or New Delhi metallo-beta-lactamase (NDM) or exhibit resistance via non-carbapenemase mediated pathways [3,4].

When a carbapenemase is present, bacteria are considered resistant to all carbapenems. However, non-carbapenemase-producing strains may be carbapenem susceptibility discordant Enterobacterales (CSD-E) [5–8]. Indeed, the Centers for Disease Control and Prevention (CDC) definition of CRE was modified to include non-carbapenemase-producing CRE (non-CP CRE), including CSD-E that are ertapenem resistant but susceptible to meropenem [3, 9]. These non-CP CRE are an important classification of CRE, and recent data have sought to differentiate their underlying resistance mechanisms and clinical outcomes [10–12]. Clinical outcomes between CP CRE and non-CP CRE have shown to be relatively similar; therefore, advanced understanding of risk factors and mechanisms of potentially precursory CSD-E is clinically warranted.

Research from MD Anderson Cancer Center (MDACC) demonstrated that most CRE were non-CP CRE emerging from an ESBL-positive Enterobacterales background via changes in ESBL gene copy number variants coupled with reduced expression of outer membrane porin genes [13,14]. Other studies have identified increasing rates of ertapenem-mono-resistant isolates, demonstrating that these isolates were less likely to have carbapenemase genes upon genetic testing compared to other CRE isolates [4, 8, 11, 15]. These non-CP CRE carbapenem discordant isolates, which are increasing at MDACC, may phenotypically display ertapenem non-susceptibility and meropenem susceptibility. Furthermore, patients with leukemia and hematopoietic stem cell transplantation (HSCT) recipients may experience higher rates of non-CP CSD-E due to extensive antibiotic exposure, complicating the prevention or treatment of infections caused by these organisms. Black et al. identified that ertapenem-resistant/meropenem-susceptible Enterobacterales had on average 4-fold more CTX-M copies than CTX-M-positive but ertapenem-susceptible/meropenem-susceptible strains for both *Escherichia coli* and *Klebsiella pneumoniae*, regardless of carbapenemase status [8]. As increased CTX-M production and loss of outer membrane porins (e.g., *omp*C) are critical for the development of the ertapenem-resistant/meropenem-susceptible phenotype in Enterobacterales, it is possible that Enterobacterales isolates with this phenotype are a precursor to the fully carbapenem-resistant non-CP Enterobacterales isolates observed at MDACC and elsewhere.

There is a paucity of literature on clinical risk factors that lead to CSD-E development in immunocompromised patients, particularly those with prior Enterobacterales infection. This literature gap is relevant as both CP-CRE and non-CP CRE pose a high burden in immunocompromised patients, including increased mortality [6, 16]. Given limited clinical experience in immunocompromised patients, we aimed to characterize risk factors for CSD-E BSI development in patients with cancer.

## Materials and Methods

### Study Design, Patient Selection, Definitions, Data Collection, Antibiotic Use Assessment, and Objectives

This is a retrospective, single-center matched case-control study comparing adults ≥ 18 years of age from January 2016 through November 2023 with a history of Enterobacterales BSI. We included patients with ≥ 2 BSIs having the same organism within one year which were either initial carbapenem susceptibility concordant (CSC)-E BSI to subsequent CSC-E BSI or initial CSC-E BSI to subsequent CSD-E BSI. CSC-E BSI isolates were ertapenem and meropenem susceptible. We chose relapses after initial infection to isolate risk factors occurring between the two episodes that may have led to CSD-E BSI. Patients were excluded if they had multiple gram-negative pathogens isolated from the blood culture of interest, meropenem-resistant Enterobacterales, or subsequent BSI occurring less than 10 days or greater than 365 days. We matched patients that progressed to CSD-E BSI to those that did not (1:3), based on organism. The University of Texas MDACC Institutional Review Board reviewed and approved this study.

CSD-E BSI was defined as bacteremia with a blood culture positive for an Enterobacterales isolate susceptible to meropenem but non-susceptible to ertapenem. CSC-E BSI was defined as bacteremia with a blood culture positive for an Enterobacterales isolate that was susceptible to both meropenem and ertapenem. To limit inadvertently capturing CP-CRE in the cohort, patients with isolates phenotypically resistant to both ertapenem and meropenem were excluded. The timeframe from positive initial blood culture to subsequent positive blood culture was limited between 10 to 365 days. Extended-spectrum (ES) cephalosporin was defined as cefpodoxime, ceftriaxone, ceftazidime, or cefepime.

Patient demographics, laboratory values, and antibiotic receipt were collected from the EHR. Laboratory values were collected as a mean value between the initial isolate date and subsequent isolate of interest date, including albumin (g/dL), weight (kg), creatine clearance [CrCl (mL/min)], and absolute neutrophil count [ANC (k/uL)]. We collected primary malignancy, HSCT recipient (yes/no), identified organism, patient death date, days between subsequent isolate of interest and death, inpatient length of stay (days), intensive care unit (ICU) admission, ICU length of stay (days), days of neutropenia, and degree of immunosuppression (high/low). Patients were considered highly immunosuppressed if their underlying malignancy diagnosis was acute myeloid leukemia (AML), acute lymphoblastic leukemia (ALL), high-risk myelodysplastic syndrome (HR-MDS), or primary hemophagocytic lymphohistiocytosis (HLH). Patients were not considered to be highly immunosuppressed if their underlying malignancy diagnosis was chronic myeloid leukemia (CML), chronic lymphocytic leukemia (CLL), aplastic anemia (AA), lymphoma, myeloma, or solid tumor(s). Neutropenia was defined as an ANC < 500 k/uL. Augmented renal clearance was defined as creatinine clearance (CrCl) ≥ 130 ml/min.

An antibiotic day of therapy was defined as a calendar day on which an antibiotic was administered to the patient. Antibiotic day(s) of therapy was reported regardless of the dosing frequency. Antibiotic use was assessed for each antibiotic and reported as a binary variable for exposure and cumulative exposure in days between the dates of the initial blood culture isolate and the subsequent blood culture isolate. Antibiotics identified for data collection included older agents (aztreonam, cefpodoxime, cefepime, ceftriaxone, ceftazidime, ciprofloxacin, levofloxacin, piperacillin-tazobactam, meropenem, ertapenem) and novel agents (ceftazidime-avibactam, meropenem-vaborbactam, ceftolozane-tazobactam, cefiderocol).

Susceptibility testing at time of infection was performed in accordance with routine hospital practice and Clinical Laboratory and Standards Institute (CLSI) guidelines [17] using a combination of Vitek2 (bioMérieux; Marcy-L’Étoile, France), BCID2 (bioMérieux; Marcy-L’Étoile, France), Accelerate Pheno (Accelerate Diagnostics; Tuscon, AZ), and/or gradient strip testing.

### Genomic Analysis

Sequencing data from serial Enterobacterales isolates has been described previously [14, 18, 19]. MLST was assigned using the PubMLST database [20]. Pairwise single nucleotide polymorphism (SNP) distances between each respective serial isolate was ascertained using the Snippy-v4.6.0 variant calling pipeline [GitHub, Seemann T, https://github.com/tseemann/snippy]. Furthermore, *ompC* and *ompF* nonsense and frameshift mutations were assessed with Snippy output. Lastly, the COpy Number Variant QuantifICation Tool [Convict; GitHub, Shropshire W, https://github.com/wshropshire/convict] was used to identify and quantify ESBL gene copy number estimates in each respective isolate and assess potential fold change in copy numbers. Relapsing serial isolates were defined as having ≤ 15 SNPs per previous definitions of genetic relatedness thresholds for Enterobacterales [21]. To determine the presence of beta-lactam survival mechanisms (BLSM) and if CSD-E serial isolates are a possible precursor to full carbapenem resistance in non-CP-CRE, serial isolate pairs underwent meropenem modified Kirby-Bauer disk diffusion testing (TDTest) using CLSI susceptibility interpretive criteria [17, 22]. Meropenem heteroresistance was defined as isolates with CLSI interpretation of intermediate or susceptible with growth of 5 or more bacterial colonies in the zone of inhibition after 24-hour antibiotic exposure. Meropenem tolerance was defined as isolates with CLSI interpretation of intermediate or susceptible with growth of 5 or more bacterial colonies in the zone of inhibition after the addition of glucose and casamino acids after 48-hour antibiotic exposure. Subsequently, bacterial colonies detected within the zone of inhibition for serial isolates underwent PAP to further evaluate meropenem heteroresistance, which we defined as the proportion of total colonies at ≥ 10^-6^ at the CLSI meropenem susceptible breakpoint (1 µg/mL) [23].

### Objectives

The primary objective was to characterize factors associated with an increased risk of CSD-E BSI following initial CSC-E BSI. Exploratory secondary objectives were the following: most prevalent CSD-E isolate species observed, definitive therapy against CSD-E, utilization of novel agents before CSD-E development, all-cause 14 and 30-day mortality of CSD-E BSI compared to CSC-E BSI, number of subsequent BSIs following CSD-E BSI, genomic and BLSM characterization of available CSD-E and CSC-E isolates.

### Statistical Analysis

Our study summarized patient characteristics using median (range) for continuous variables and frequencies (percentages) for categorical variables. Fisher’s exact test was used to compare categorical variables while the Wilcoxon rank-sum test was employed for continuous variables.

Propensity score matching (PSM) was used to match subjects, balancing the distribution of the organism variable between case and control cohorts. Matching was implemented using the “MatchIt” package in R. Logistic regression models were employed to evaluate risk factors associated with the CSD-E isolate, including a binary antibiotic exposure variable.

Time-varying covariate Cox proportional hazards models were used to account for time-dependent changes in antibiotic exposure when estimating the association between antibiotic use and CD event. Patients who died before experiencing CSD-E were censored at time of death. The time-varying Cox proportional hazards model accounts for time bias and allows for an assessment of risk of coalescence associated with each additional day of antibiotic exposure. Univariate models were fitted, and factors (cumulative antibiotic use) with p values ≤0.2 in these models were included in the initial multivariable model. The final multivariable model was obtained through backward elimination, retaining only factors with p values ≤0.2. All statistical analyses were conducted in R (version 4.4.1) and statistical significance was achieved at p=0.05.

## Results

We evaluated 829 patients with Enterobacterales BSI (Figure 1). Repeat BSI was CSC-E in 81 patients (9.8%) and CSD-E in 14 patients (1.7%). Twelve patients with repeat CSD-E BSI had an initial ceftriaxone resistant Enterobacterales (CRO-R-E) isolate (85.7%) within the prior year. Seventy-five patients with repeat CSC-E BSI had an initial CRO-R-E isolate (92.6%) within the prior year (Figure 1).

**Figure 1:**
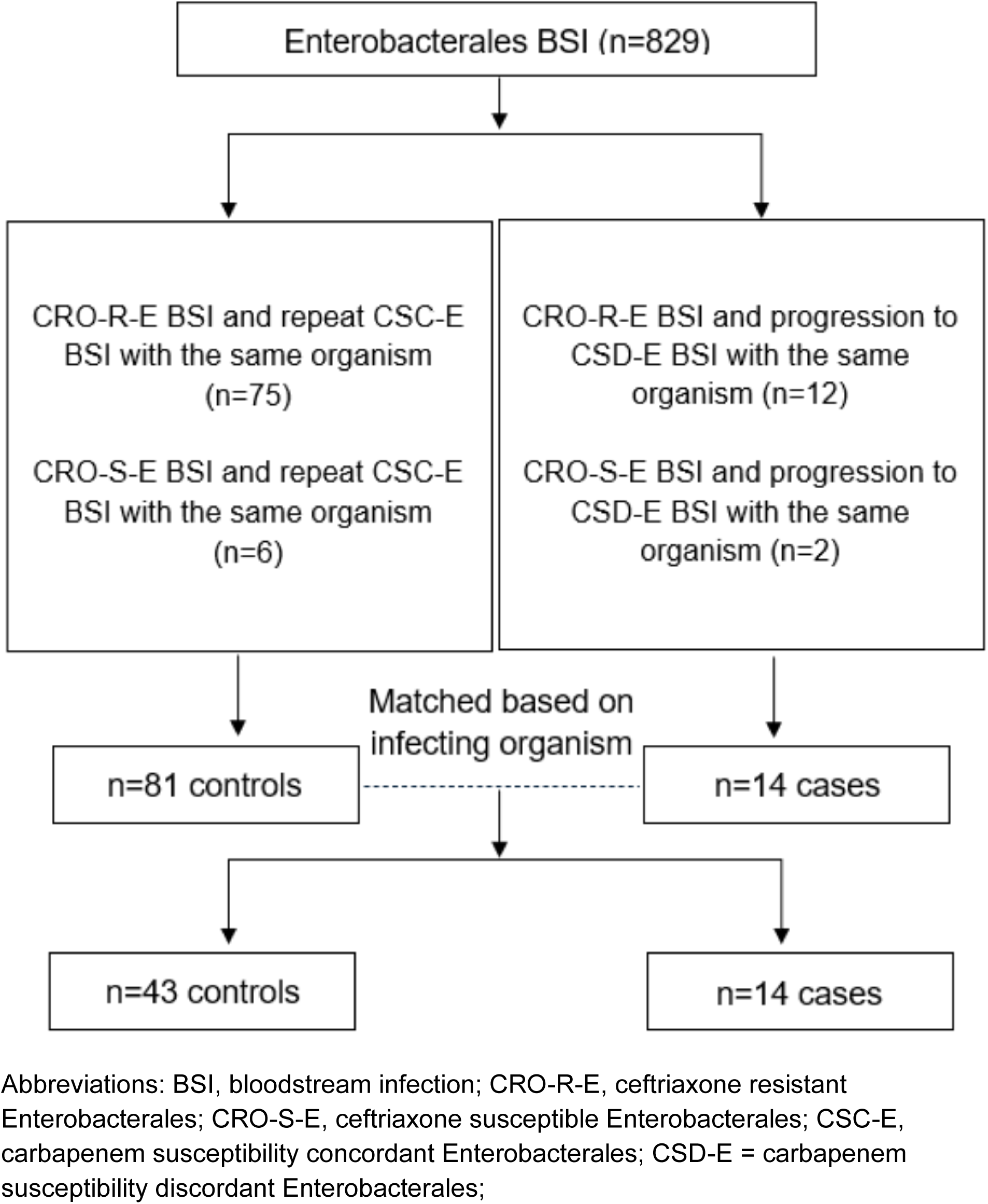
Patient Distribution and Matching

After matching, there were 43 CSC-E controls and 14 CSD-E cases. *Escherichia coli* (n=7, 50.0%), *Klebsiella* species [*Klebsiella pneumoniae,* n=5 (35.7%); *Klebsiella oxytoca,* n=1 (7.1%)], and *Enterobacter cloacae* (n=1, 7.1%) were the most common organisms in the case cohort. *Escherichia coli* (n=30, 69.8%), *Klebsiella* species [*Klebsiella pneumoniae,* n=10 (23.2%)], and *Enterobacter cloacae* (n=3, 7.0%) were the most common organisms in the control cohort. Baseline demographics are shown in Table 1. There were no differences in age, sex, or ethnicity. CSC-E and CSD-E patients were primarily classified as highly immunosuppressed (n=34, 79.1% and n=12, 85.7%, respectively). Subsequent CSC-E or CSD-E BSI occurred most often within 90 days of the initial BSI (n=35, 81.4% and n=12, 85.7%, respectively).

**Table 1.**
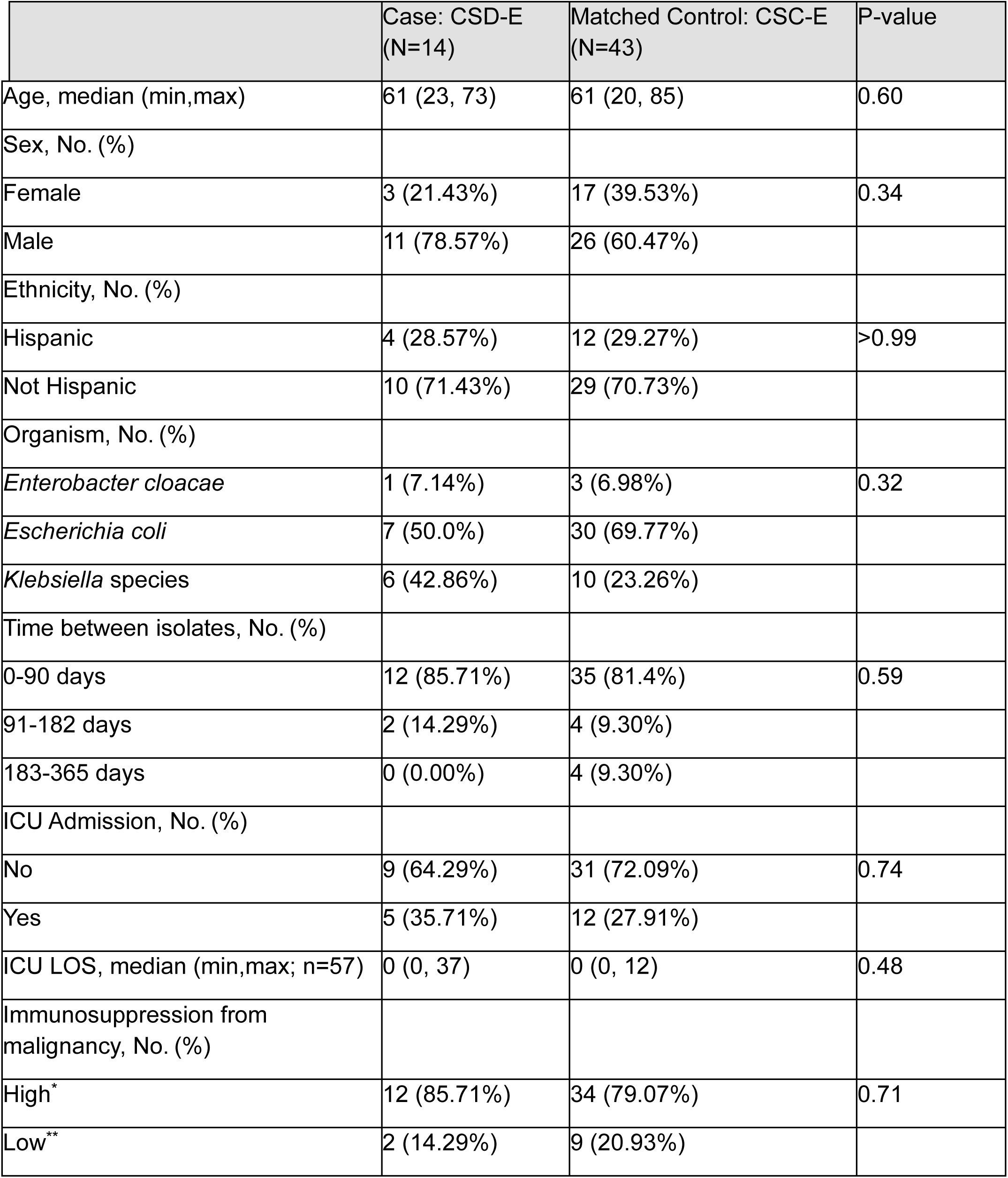
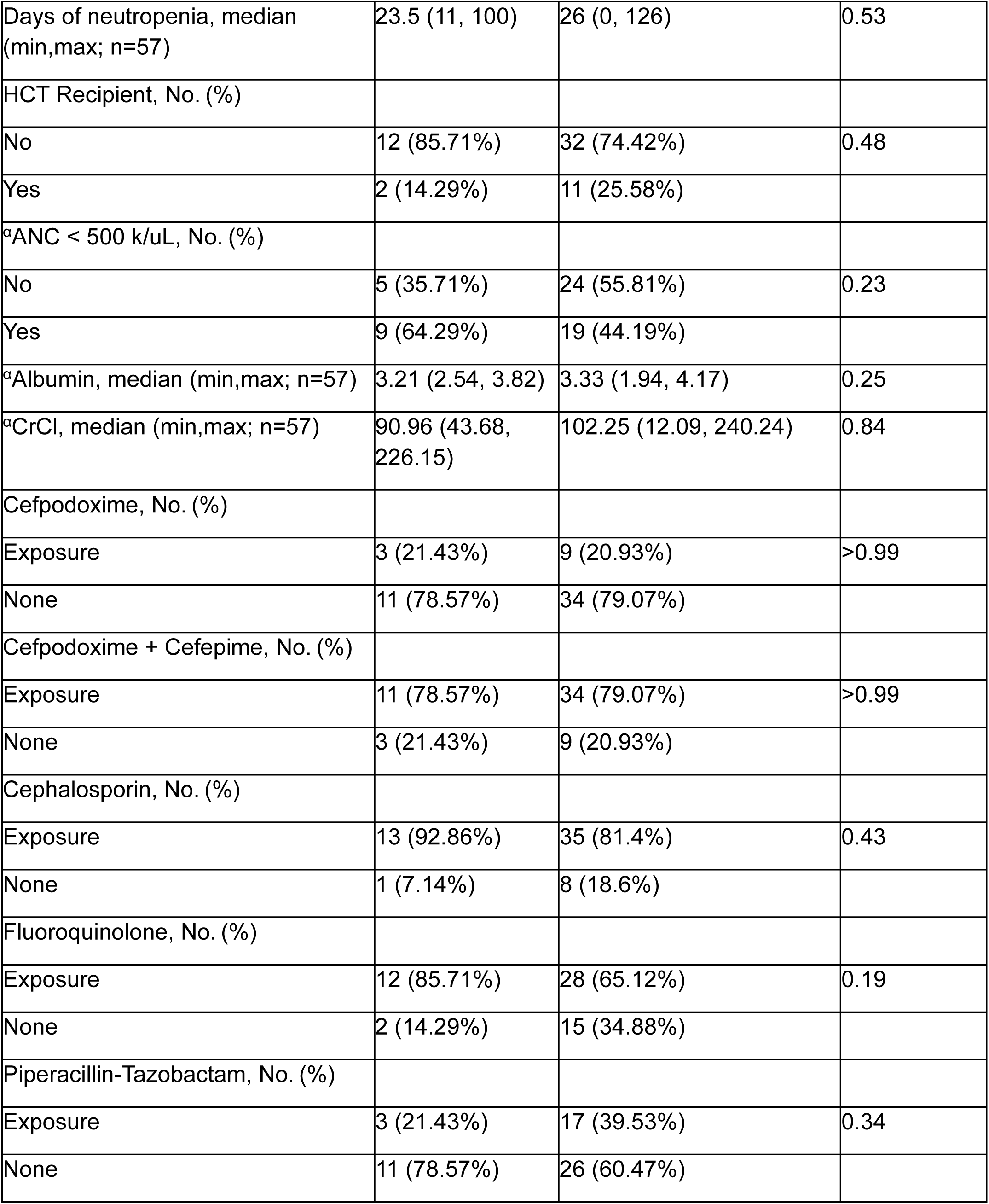
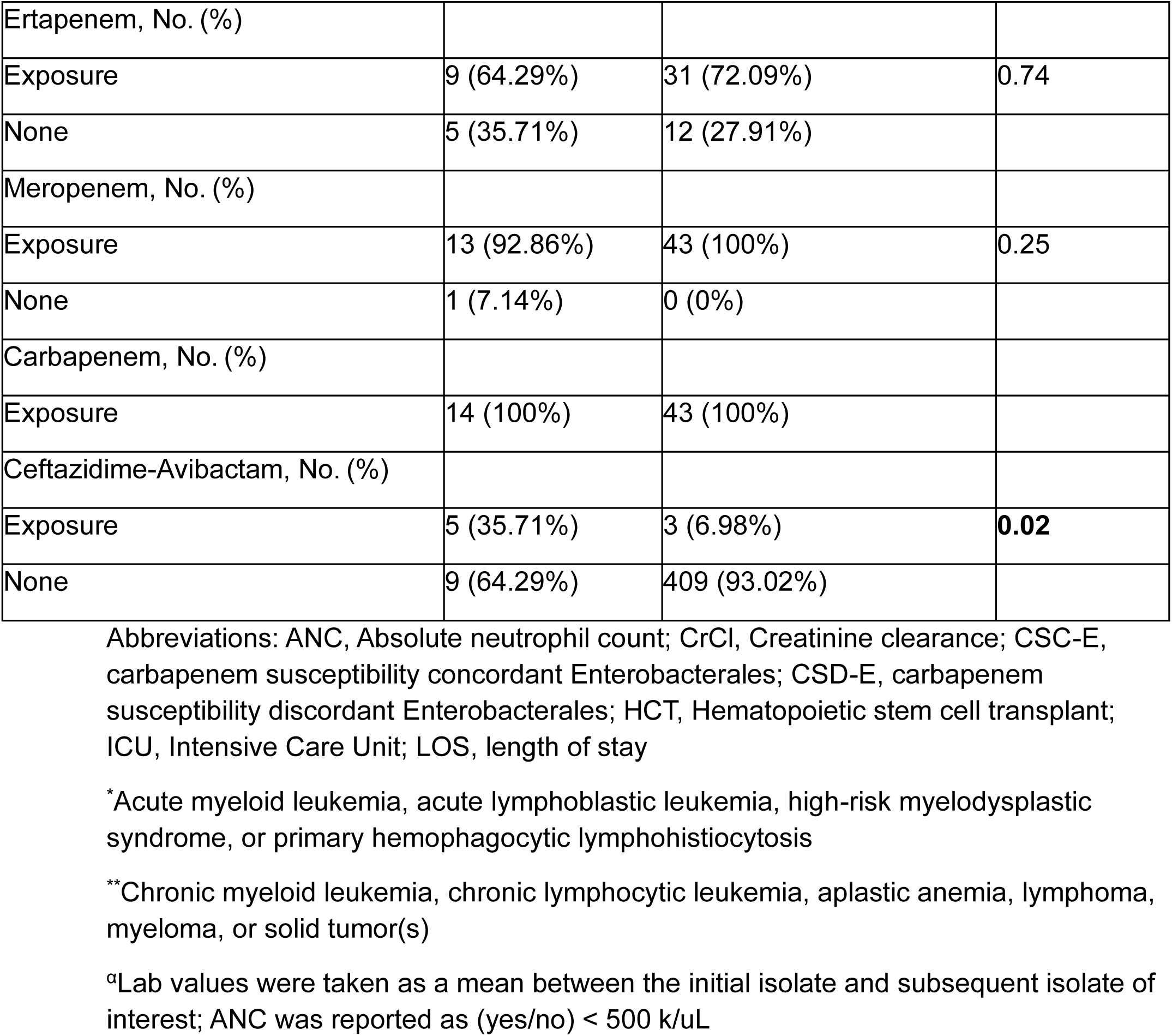
Baseline Demographics of Cases and Matched Controls.

In univariate logistic regression analysis, any CZA exposure was significantly associated with developing CSD-E BSI (odds ratio [OR], 7.41 p=0.01) (Table 2). Highly immunosuppressed patients and patients with an ANC < 500 k/uL had higher odds of developing a CSD-E BSI, though not statistically significant (OR, 1.59; p=0.59 and OR, 2.27, p=0.20, respectively). In the univariate analysis, time-varying covariate Cox regression identified each additional day of cefepime to be significantly associated with CSD-E BSI development (HR, 1.054; p=0.018); furthermore, each additional day of ceftazidime and meropenem trended towards significant association with CSD-E BSI development (HR, 1.079; p=0.07 and HR, 1.052; p=0.07, respectively) (Table 2; Figure 2).

**Figure 2:**
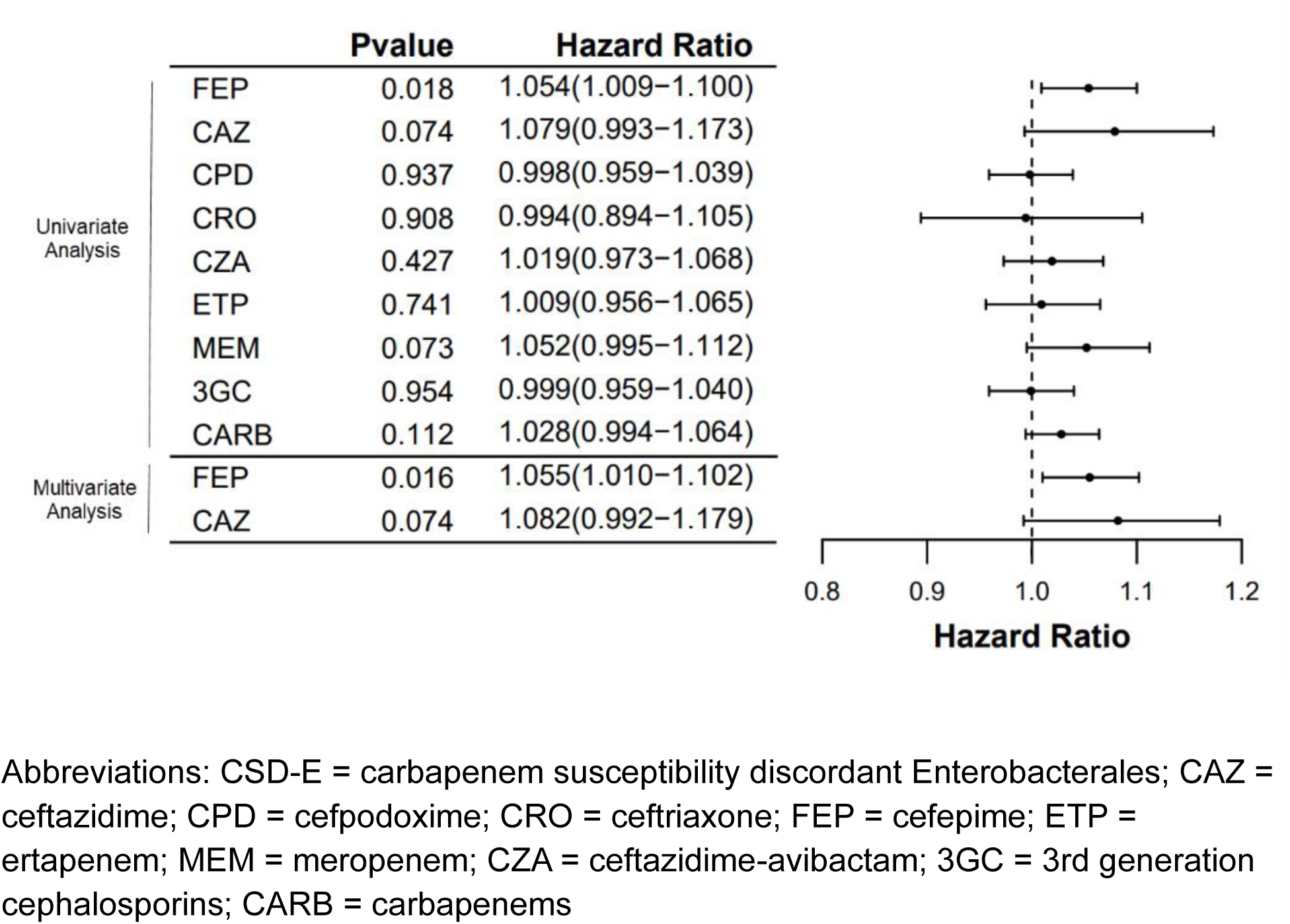
Time-varying Cumulative Antibiotic Exposure and CSD-E Risk

**Table 2.** Univariate Analysis of Cases and Matched Controls.

| Patient Demographics |  | Odds ratio | OR 95% CI | P-value |
| --- | --- | --- | --- | --- |
| Age |  | 0.99 | (0.96-1.03) | 0.70 |
| Gender | Female versus Male | 0.42 | (0.1-1.71) | 0.23 |
| Ethnicity | Not Hispanic versus Hispanic | 1.03 | (0.27-3.92) | 0.96 |
| ICU Admission | Yes versus No | 1.44 | (0.4-5.13) | 0.58 |
| HCT Recipient | Yes versus No | 0.48 | (0.09-2.52) | 0.39 |
| ICU LOS |  | 1.07 | (0.97-1.18) | 0.19 |
| °Albumin |  | 0.55 | (0.15-1.96) | 0.35 |
| °CrCl |  | 1 | (0.98-1.02) | 0.60 |
| °ANC < 500 k/uL | Yes versus No | 2.27 | (0.65-7.97) | 0.20 |
| Time between isolates | 91-182 days versus 0-90 days | 1.46 | (0.24-9.03) | 0.68 |
|  | 183-365 days versus 0-90 days | 0 | (0-Inf) | 0.99 |
| Days of Neutropenia |  | 1 | (0.98-1.02) | 0.87 |
| Immunosuppression from malignancy | High* vs Low** | 1.59 | (0.3-8.4) | 0.59 |
| Organism | *Enterobacter cloacae* versus *Escherichia coli* | 1.43 | (0.13-15.92) | 0.77 |
|  | *Klebsiella* species versus *Escherichia coli* | 2.57 | (0.69-9.56) | 0.16 |
| Antibiotic Exposure |  | Odds ratio | OR 95% CI | P-value |
| Cefpodoxime | Exposure versus None | 1.03 | (0.24-4.48) | 0.97 |
| Cefpodoxime + Cefepime | Exposure versus None | 0.97 | (0.22-4.22) | 0.97 |
| Cephalosporin | Exposure versus None | 2.97 | (0.34-26.17) | 0.33 |
| Fluoroquinolone | Exposure versus None | 3.21 | (0.63-16.35) | 0.16 |
| Piperacillin-Tazobactam | Exposure versus None | 0.42 | (0.1-1.71) | 0.23 |
| Ertapenem | Exposure versus None | 0.7 | (0.19-2.49) | 0.58 |
| Meropenem | Exposure versus None | 0 | (0-Inf) | 0.99 |
| Ceftazidime-Avibactam | Exposure versus None | 7.41 | (1.48-36.95) | **0.01** |
| Antibiotic Exposure |  | Hazard Ratio | HR 95% CI | P-value |
| Cefpodoxime | Cumulative Exposure | 0.998 | 0.959 - 1.039 | 0.937 |
| Ceftriaxone | Cumulative Exposure | 0.994 | 0.894 - 1.105 | 0.908 |
| Ceftazidime | Cumulative Exposure | 1.079 | 0.993 - 1.173 | 0.074 |
| Cefepime | Cumulative Exposure | 1.054 | 1.009 - 1.100 | **0.018** |
| Ertapenem | Cumulative Exposure | 1.009 | 0.956 - 1.065 | 0.741 |
| Meropenem | Cumulative Exposure | 1.052 | 0.995 - 1.112 | 0.073 |
| Ceftazidime-Avibactam | Cumulative Exposure | 1.019 | 0.973 - 1.068 | 0.427 |
| 3GC*** | Cumulative Exposure | 0.999 | 0.959 - 1.040 | 0.954 |
| Carbapenem | Cumulative Exposure | 1.028 | 0.994 - 1.064 | 0.112 |

Abbreviations: ANC, Absolute neutrophil count; CrCl, Creatinine clearance; CSD-E, Carbapenem susceptibility discordant Enterobacterales; HCT, Hematopoietic stem cell transplant; ICU, Intensive Care Unit; LOS, length of stay
\*Acute myeloid leukemia, acute lymphoblastic leukemia, high-risk myelodysplastic syndrome, or primary hemophagocytic lymphohistiocytosis
\*\*Chronic myeloid leukemia, chronic lymphocytic leukemia, aplastic anemia, lymphoma, myeloma, or solid tumor(s)
\*\*\*3GC (third generation cephalosporins): cefpodoxime, ceftriaxone, ceftazidime
<sup>a</sup>Lab values were taken as a mean between the initial isolate and subsequent isolate of interest; ANC was reported as (yes/no) < 500 k/uL

In the multivariable analysis, each additional day of cefepime remained significantly associated with an increased risk of CSD-E BSI development (HR, 1.055; p=0.016), whereas ceftazidime trended towards significance (HR, 1.082; p=0.07) (Table 3).

**Table 3.** Exploratory Multivariable Analysis and CSD-E Risk.

|  | Hazard Ratio | HR 95% CI | P-value |
| --- | --- | --- | --- |
| Ceftazidime cumulative | 1.082 | 0.992 - 1.179 | 0.074 |
| Cefepime cumulative | 1.055 | 1.010 - 1.102 | **0.016** |

Abbreviations: CSD-E, carbapenem susceptibility discordant Enterobacterales

In both univariate and multivariable analyses, no other identified beta-lactam or fluoroquinolone was significantly correlated with CSD-E BSI development when considering overall exposure and cumulative exposure. Mean albumin, mean CrCl, and mean ANC were not significantly correlated with CSD-E BSI development.

Following CSD-E BSI occurrence, meropenem was the primary definitive treatment of choice (n=7, 50.0%) followed by CZA (n=5, 35.7%). Of the patients in the case cohort that received CZA or meropenem as definitive treatment (n=12, 85.7%), patients received a median of 15 days of CZA and 20 days of meropenem. Eleven patients in the case cohort (78.6%) died with median time to death of 29 days following CSD-E BSI. Mortality within 14 days occurred in 1 case and 3 matched controls, with crude mortality rates of 7.1% and 6.98%, respectively. Mortality within 30 days occurred in 6 cases and 10 matched controls, with crude mortality rates of 42.9% and 23.2%, respectively. There were no significant differences in 14- or 30-day mortality between the cases and matched controls. Of the 14 CSD-E BSI, 4 patients (28.6%) developed another BSI with the same organism in the following year (n=2, 50% had ertapenem and meropenem resistant phenotype; n=2, 50% had ertapenem and meropenem susceptible phenotype).

Seven patients (50.0%) in the case cohort and 11 patients (25.6%) in the matched control cohort had genomic data available for both the initial isolate and subsequent isolate of interest, yielding 14 and 22 historical isolates available for analysis, respectively. Of the 7 patients in the case cohort with genomic data available, 4 patients had BSI caused by *E. coli*, and 3 had BSI caused by *K. pneumoniae.* All subsequent case isolates were of the same sequence type as initial case isolates and considered relapsed infection versus reinfection. Case 1 demonstrated a 5-fold and 6-fold increase in CTX-M and OXA-1 copy number variants (CNVs), respectively, with TDTest revealing observable meropenem heteroresistant subpopulations within the zone of inhibition at 24 hours. Furthermore, PAP revealed the overall fraction of the population increases at higher meropenem concentration for the serial CSD-E isolate, consistent with borderline meropenem heteroresistance (Table 4; Supplemental Figure 2). Cases 2 and 6 demonstrated no appreciable beta-lactamase CNV changes but harbored an *omp*K36 and *omp*F mutation, respectively. Case 3 demonstrated a >2-fold increase in TEM CNVs. Case 7 demonstrated a 6-fold increase in TEM CNVs, coupled with an *omp*F mutation. Cases 4 and 5 did not demonstrate increased beta-lactamase CNV changes or *omp* mutations; however, case 5 TDTest revealed observable meropenem tolerant subpopulations within the zone of inhibition at 48 hours and subsequent PAP evaluation was consistent with a meropenem tolerant phenotype. Two matched control patients had isolates with mismatching sequence types consistent with reinfection and were excluded. Of the remaining 9 patients in the matched control cohort, 4 patients had BSI caused by *E. coli*, and 5 had BSI caused by *K. pneumoniae*. No matched control isolates demonstrated a >2-fold CNV increase in any beta-lactamase or Omp mutations from the initial isolate to the subsequent isolate. BLSM were not observed in the matched control isolates. We evaluated the identification of recurrent BSI by measuring pairwise SNP distances and found less than 15 SNPs for all case isolates, confirming beta-lactamase amplification and reduced Omp expression are possible mechanisms implicated in the development of CSD-E BSI in these cases.

**Table 4.**
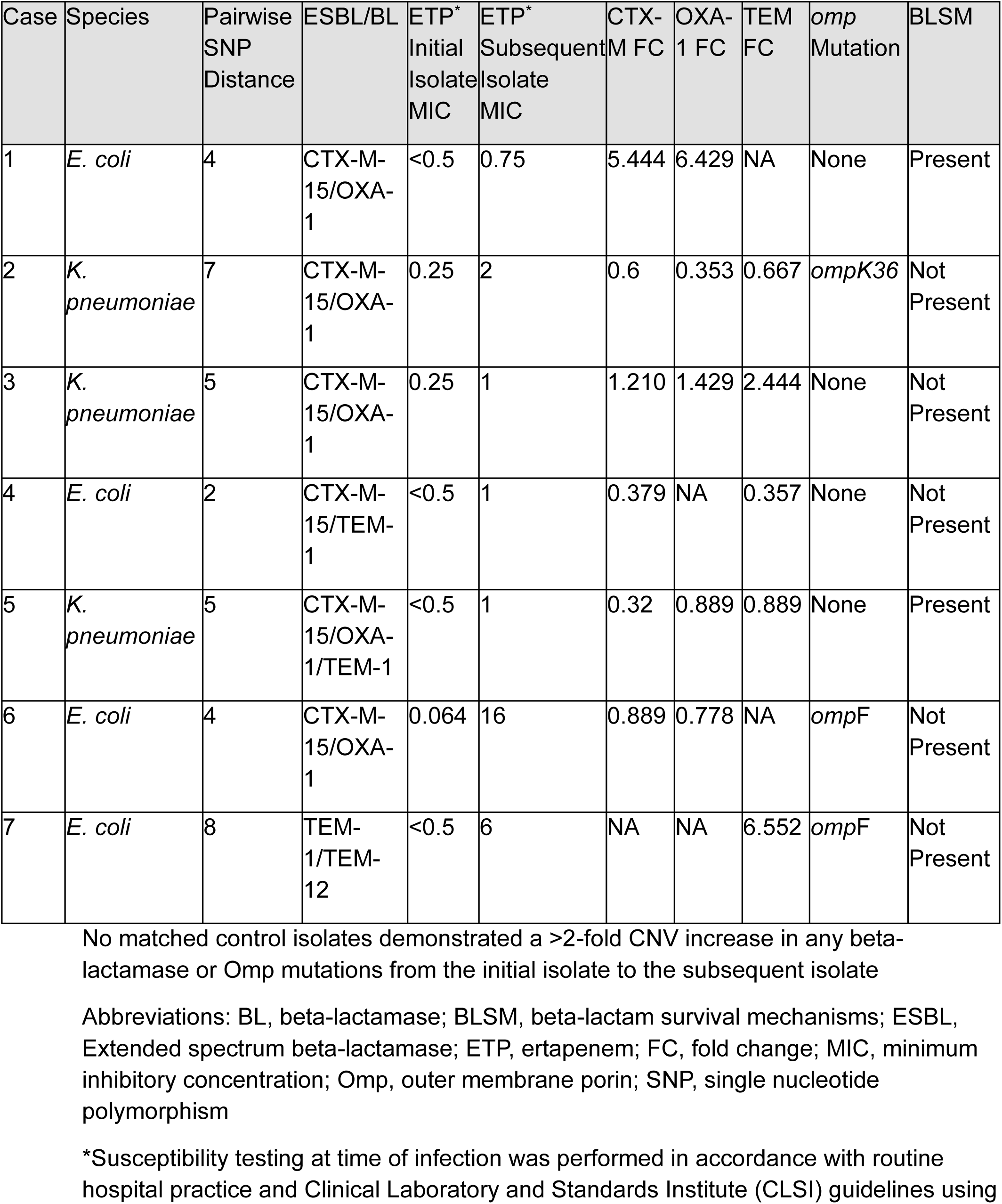
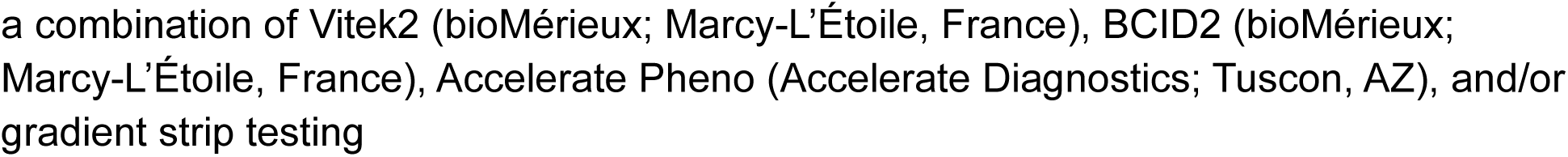
CSD-E Genomic Analysis.

| Case | Species | Pairwise<br>SNP<br>Distance | ESBL/BL | ETP*<br>Initial<br>Isolate<br>MIC | ETP*<br>Subsequent<br>Isolate<br>MIC | CTX-M FC | OXA-1 FC | TEM<br>FC | omp<br>Mutation | BLSM |
| --- | --- | --- | --- | --- | --- | --- | --- | --- | --- | --- |
| 1 | *E. coli* | 4 | CTX-M-15/OXA-1 | <0.5 | 0.75 | 5.444 | 6.429 | NA | None | Present |
| 2 | *K. pneumoniae* | 7 | CTX-M-15/OXA-1 | 0.25 | 2 | 0.6 | 0.353 | 0.667 | ompK36 | Not<br>Present |
| 3 | *K. pneumoniae* | 5 | CTX-M-15/OXA-1 | 0.25 | 1 | 1.210 | 1.429 | 2.444 | None | Not<br>Present |
| 4 | *E. coli* | 2 | CTX-M-15/TEM-1 | <0.5 | 1 | 0.379 | NA | 0.357 | None | Not<br>Present |
| 5 | *K. pneumoniae* | 5 | CTX-M-15/OXA-1/TEM-1 | <0.5 | 1 | 0.32 | 0.889 | 0.889 | None | Present |
| 6 | *E. coli* | 4 | CTX-M-15/OXA-1 | 0.064 | 16 | 0.889 | 0.778 | NA | ompF | Not<br>Present |
| 7 | *E. coli* | 8 | TEM-1/TEM-12 | <0.5 | 6 | NA | NA | 6.552 | ompF | Not<br>Present |

No matched control isolates demonstrated a >2-fold CNV increase in any beta-lactamase or Omp mutations from the initial isolate to the subsequent isolate
Abbreviations: BL, beta-lactamase; BLSM, beta-lactam survival mechanisms; ESBL, Extended spectrum beta-lactamase; ETP, ertapenem; FC, fold change; MIC, minimum inhibitory concentration; Omp, outer membrane porin; SNP, single nucleotide polymorphism
\*Susceptibility testing at time of infection was performed in accordance with routine hospital practice and Clinical Laboratory and Standards Institute (CLSI) guidelines using
a combination of Vitek2 (bioMérieux; Marcy-L'Étoile, France), BCID2 (bioMérieux; Marcy-L'Étoile, France), Accelerate Pheno (Accelerate Diagnostics; Tuscon, AZ), and/or gradient strip testing

## Discussion

Patient-specific risk factors for CSD-E infection have been previously described for the general population [4]. Our study examines risk factors, including antibiotic exposures, for CSD-E development in immunocompromised patients. As previously described, CSD-E can emerge from an ESBL-positive Enterobacterales background, especially with concomitant porin inactivation that reduces antibiotic entry [5–8, 14]. Thus, it is feasible that cumulative cephalosporin therapy could be associated with an increase in CSD-E BSI development by selecting for these resistance pathways. Indeed, our study revealed each additional day of cefepime use significantly increased the hazard of CSD-E development by 5.5%; ceftazidime trended towards significantly increased hazard of CSD-E development by 8.2% (p=0.07) (Figure 2). These findings are plausible based on studies demonstrating variable ESBL hydrolysis rates of latter-generation cephalosporins depending on the implicated ESBL enzyme [24]. In addition, the inoculum effect on ceftazidime and cefepime has been demonstrated in CTX-M-ESBL-producing *Escherichia coli*, where the MIC of ceftazidime and cefepime increases as the bacterial inoculum increases, leading to a potential 8-fold to 64-fold increase in MIC, respectively [25]. Despite being a substrate for beta-lactamase hydrolysis, ceftriaxone did not show a significant association with CSD-E BSI development; most of the CSD-E cohort had initial CRO-R-E, and immunocompromised patients are more likely to receive broader-spectrum anti-pseudomonal agents during periods of febrile neutropenia. In our study, no significant differences were observed between the cohorts for any or cumulative exposure of meropenem or ertapenem. Furthermore, median exposure to ertapenem was higher in the control group that did not develop CSD-E BSI. Ertapenem exposure in the CSD-E group is shown in Supplemental Figure 1. Overall, understanding which antibiotics may be associated with downstream CSD-E is critical, as most patients who developed CSD-E BSI began with an initial CRO-R-E BSI yet received cefepime and/or ceftazidime before CSD-E development; this suggests a minimally appropriate amount of an active agent is needed to treat infection and avoid propagating and amplifying resistance. Comparative genomics highlighted CNV increases in beta-lactamases and/or Omp mutations for available CSD-E isolates which were not seen in the matched controls. Additionally, we hypothesize that CSD-E serial isolates may represent an evolutionary intermediate stage towards fully carbapenem-resistant non-CP Enterobacterales isolates; indeed, phenotypic characterization of isolates with available paired WGS data revealed the presence of meropenem heteroresistant and tolerant subpopulations within the zone of inhibition in 2 out of 7 CSD-E case pairs compared to none in the matched control group. Further studies are necessary to confirm if CSD-E serial isolates are at greater risk of meropenem heteroresistance or tolerance, serving as an evolutionary stage towards fully carbapenem resistant non-CP Enterobacterales.

Patients exposed to CZA had 7.41 times higher odds of developing a CSD-E BSI compared to patients with no CZA exposure when treated as a binary variable in univariate analysis. When treated as a time-varying covariate, CZA cumulative exposure was not significantly associated with CSD-E BSI development (HR, 1.019; p=0.427). Based on the resistance mechanisms leading to CSD-E BSI development, we expect that avibactam, a broad beta-lactamase inhibitor, protects ceftazidime from beta-lactamase hydrolysis, even in the setting of beta-lactamase gene amplification [26]. However, porin deletions in beta-lactamase-producing strains have shown to increase CZA MIC [27]. This may explain the observed discrepancy between any and cumulative CZA exposure for CSD-E BSI development but requires further investigation with larger cohorts. CZA was initiated in the CSD-E cohort between the initial and subsequent BSIs primarily empirically or for non-bloodstream infections with non-carbapenemase producing carbapenem resistant organisms other than Enterobacterales.

Characterizing specific risk factors for CSD-E development in immunocompromised patients is important to understand clinical outcomes; indeed, Adelman et al. describe a trend towards CSD-E development for patients with malignancy, and 2.45 times higher odds of 90-day mortality amongst patients with malignancy and CSD-E infection [4]. Recent data suggests patients with CSD-E CRE infections have similar outcomes as those with multi-carbapenem-resistant Enterobacterales; however, cancer/immune status was not reported for this subset of CRACKLE-2 patients [11, 12]. All patients in our study who developed CSD-E BSI had a primary hematologic malignancy, predominantly AML (n=10, 71.4%). Univariate analysis did not reveal significant differences among the two cohorts regarding HSCT or days of neutropenia between the initial and subsequent BSI isolates. Mean ANC < 500 k/uL (during the period between initial and subsequent isolates) and highly immunosuppressed cancer status were associated with higher odds of CSD-E development, but the findings were not statistically significant. This may be due to our small sample size, warranting further investigation in larger studies, as these factors have been implicated in resistance development and increased mortality for gram-negative bacteremia [28]. Prior studies have also demonstrated that hypoalbuminemic patients with carbapenem-susceptible Enterobacterales BSI are at 4.6 times higher odds of 30-day mortality when receiving ertapenem compared to receiving meropenem or imipenem-cilastatin [29]. This may be exacerbated in patients with augmented renal clearance where free fraction of ertapenem is more readily cleared, leading to subtherapeutic exposure, therapy failure, and possibly resistance development. The CSD-E cohort did not have a significant difference in hypoalbuminemia or CrCl (including augmented renal clearance). There were no patients with hypoalbuminemia who developed CSD-E BSI, which limited evaluation of outcomes relating to hypoalbuminemia and ertapenem exposure.

We acknowledge several limitations. First, our study population was comprised of adult patients with cancer. It is unknown if the findings of our study are applicable to other immunocompromised patient populations, thus multicenter evaluations of CSD-E BSI development are warranted for external validation. Furthermore, this single-center study was performed in the USA, limiting generalizability to other countries with different CRE epidemiology; however, only 6 patients in the case cohort (42.9%) were born in the USA. Next, these findings should be interpreted as hypothesis generating as the multivariable model may be overfit due to the small sample size. We chose a wide time frame between initial and subsequent isolates which may introduce confounders; however, repeat and surveillance blood cultures are performed frequently in our hematologic malignancy patients, increasing confidence that the 10-day minimum window would help capture recurrence and exclude persistence. Additionally, EHR functionality with synced hospital systems was leveraged to mitigate incomplete culture history and account for outpatient antibiotic prescriptions. Next, not all isolates had genomic data available, which limited definitive conclusion of which genomic mutations predominantly confer CSD-E and how they are affected by antibiotic exposure. However, we believe the combination of available genomic and clinical data for the matched isolates suggests cumulative cefepime exposure may trigger a resistance development cascade from CSC-E to CSD-E via beta-lactamase amplification and/or outer membrane porin changes, observed in 5 of 7 case isolates. This hypothesis is consistent with findings by Black et al., who demonstrated that the presence of *omp*C mutations and amplification of CTX-M is seen in CSD *E. coli* and *K. pneumoniae* strains compared to CSC-E CTX-M positive strains [8]. Furthermore, these CSD-E strains co-harbored additional beta-lactamases, such as TEM, consistent with our findings. Lastly, the physiologic source causing the BSI was not fully investigated, thus source control measures during hospitalization are unknown. However, most patients (80.7%) had a highly immunosuppressive malignancy and were neutropenic, likely leading to Enterobacterales BSI from gastrointestinal translocation due to disrupted barrier function, which is not readily amenable to source control [30–32].

Despite the limitations, we identify key strengths. Isolates were identified as early as January 2016, providing insight into the relative frequency of CSD-E BSIs and rates of recurrent CSC CRO-R-E BSIs. Each cohort included isolates from 2016 to 2023, mitigating differences in susceptibility testing, treatment strategies, and chemotherapeutic regimens across the range of years. The study was limited to bacteremia to avoid confounders of heterogenous infections, with matching performed based on organism to balance the two cohorts. Next, we expanded upon factors that may contribute to CSD-E BSI development in immunocompromised patients, including specific antibiotic exposure, enhancing clinical comprehension of how antibiotic use impacts downstream carbapenem susceptibility in Enterobacterales. To our knowledge, this is the first study evaluating the association of specific antibiotic exposure to downstream CSD-E BSI development in immunocompromised cancer patients. Additionally, incorporation of a time-varying Cox proportional hazards model assists with characterizing how cumulative antibiotic exposure may affect the risk of developing a CSD-E BSI while minimizing time bias. Finally, we also included available genomic sequencing data and BLSM evaluation to strengthen and provide mechanistic rationale for our clinical findings.

In conclusion, these data suggest that possible risk factors associated with CSD-E BSI development in immunocompromised patients include any exposure to CZA and cumulative exposure to cefepime. Further studies are warranted to validate these findings and other CSD-E BSI risk factors.

## Data Availability

All data produced in the present study are available upon reasonable request to the authors.

## Acknowledgements

This research received no specific grant from any funding agency in the public, commercial, or not-for-profit sectors. The authors declared no conflicts of interest regarding the research, authorship, and/or publication of this article.

---

**Supplemental Figure 1:**
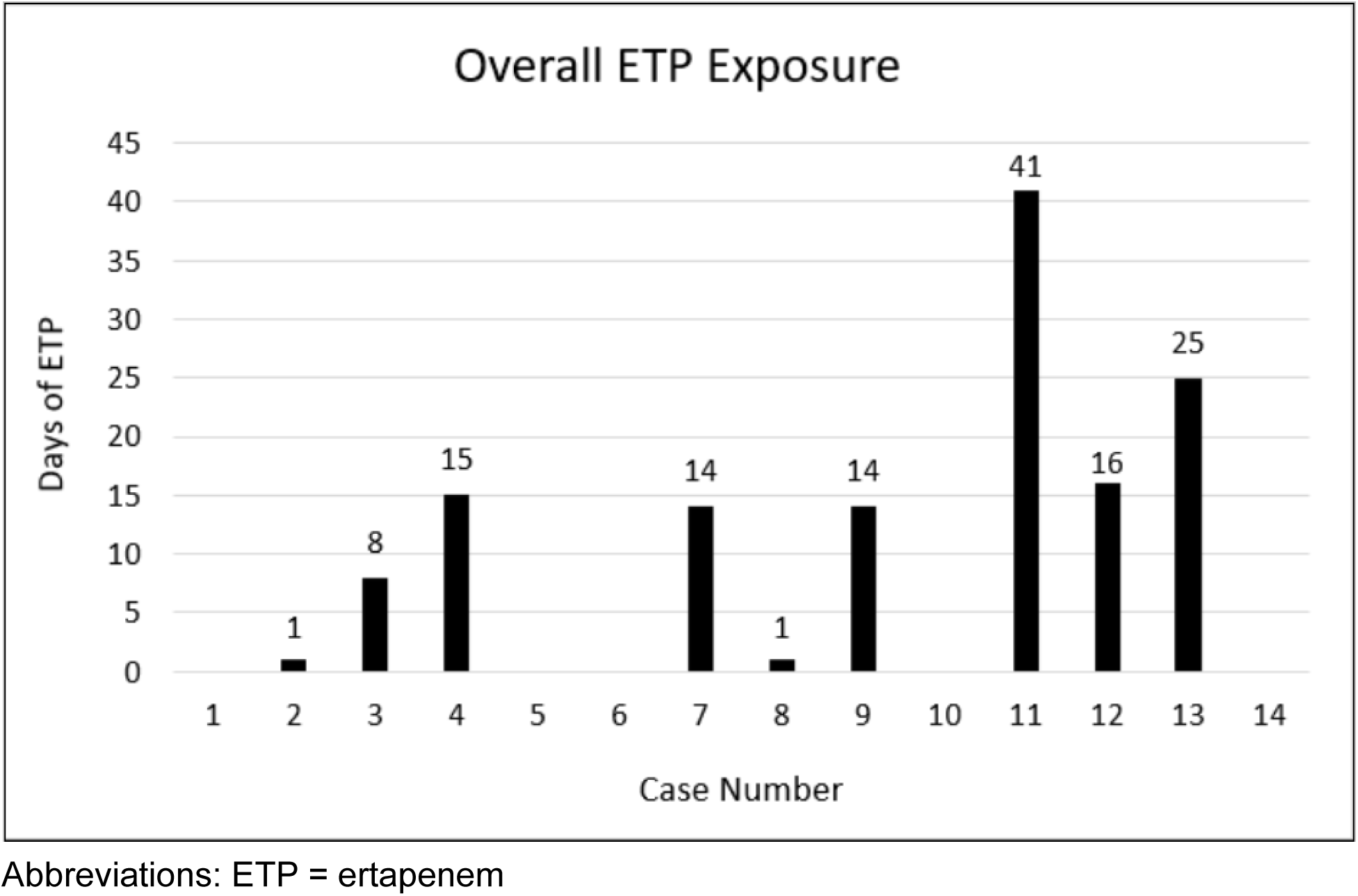
Overall Ertapenem Exposure in the Case Cohort

**Supplemental Figure 2:**
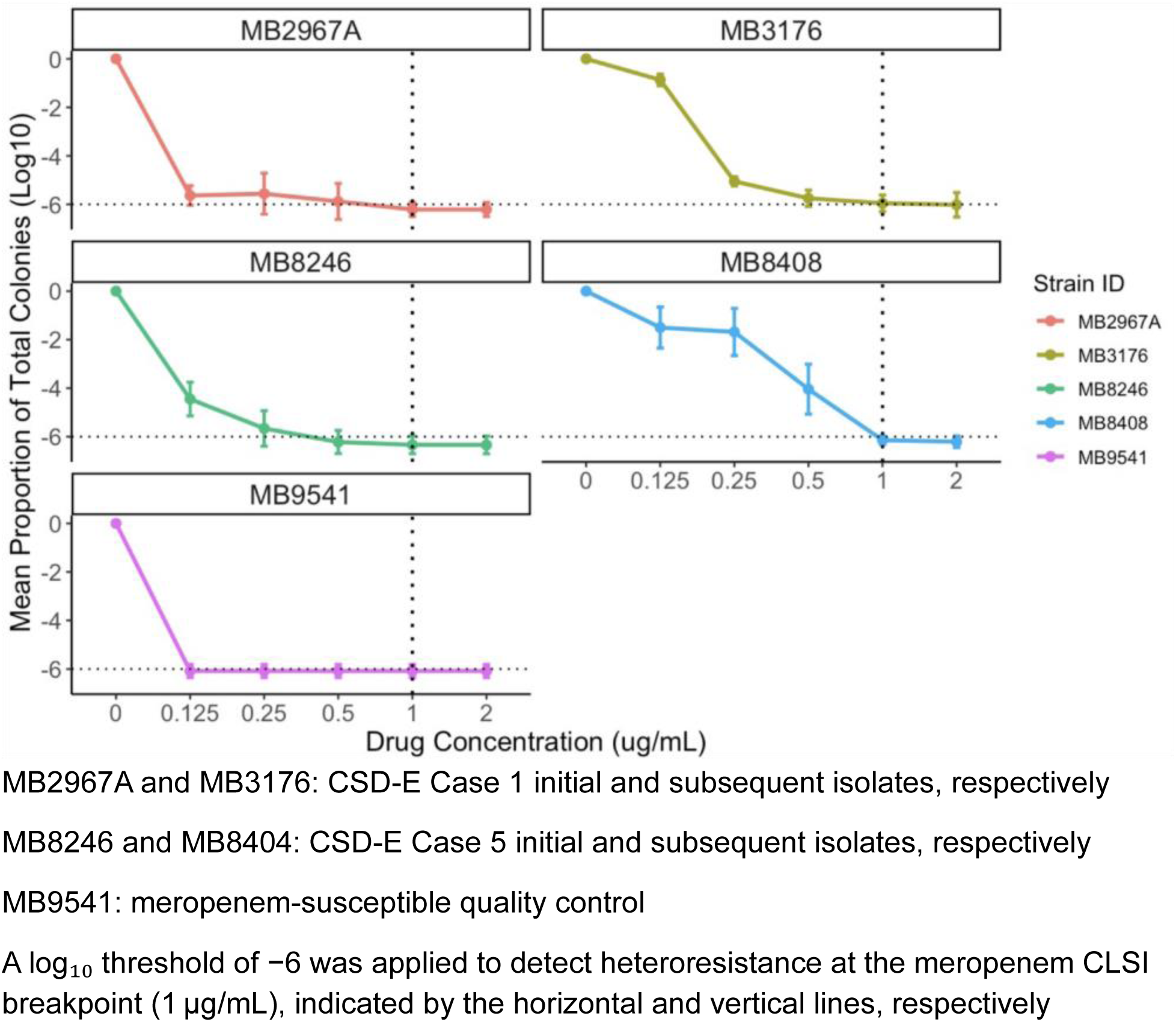
Population Analysis Profiling (PAP) to Evaluate Meropenem Heteroresistance/Tolerance:

